# Model-based estimation of transmissibility and reinfection of SARS-CoV-2 P.1 variant

**DOI:** 10.1101/2021.03.03.21252706

**Authors:** Renato Mendes Coutinho, Flavia Maria Darcie Marquitti, Leonardo Souto Ferreira, Marcelo Eduardo Borges, Rafael Lopes Paixão da Silva, Otavio Canton, Tatiana P. Portella, Silas Poloni, Caroline Franco, Mateusz M. Plucinski, Fernanda C. Lessa, Antônio Augusto Moura da Silva, Roberto Andre Kraenkel, Maria Amélia de Sousa Mascena Veras, Paulo Inácio Prado

**Author notes:** RMC, FMDM, LSF, MEB, RLPS, TPP, SP, CF, RAK, MASMV, and PIP designed research; RMC, FMDM, LSF, MEB, RLPS, TPP, SP, and PIP performed research and analyzed data; and all authors wrote the paper.

## Abstract

The variant of concern (VOC) P.1 emerged in the Amazonas state (Brazil) in November-2020. It contains a constellation of mutations, ten of them in the spike protein. Consequences of these specific mutations at the population level have been little studied so far, despite the detection of P.1 variant in 26 countries, with local transmission in at least four other countries in the Americas and Europe. Here, we estimate P.1’s transmissibility and reinfection using a model-based approach, by fitting data from the Brazilian national health surveillance of hospitalized individuals and frequency of the P.1 variant in Manaus from December 2020 to February 2021, when the city was devastated by four times more cases than in the previous peak (April 2020). The new variant was found to be about 2.6 times more transmissible (95% Confidence Interval (CI): 2.4–2.8) than previous circulating variant(s). The city already had a high prevalence of individuals previously affected by the SARS-CoV-2 virus (estimated as 78%, CI:73–83%), and the fitted model attributed 28% of the cases during the period to reinfections by the variant P.1. Our estimates rank P.1 as the most transmissible among the current identified SARS-CoV-2 VOCs, posing a serious threat and requiring urgent measures to control its global spread.

---

**T**he Japanese National Institute of Infectious Diseases identified the new P.1 SARS-CoV-2 variant from travelers returning from Amazonas State, Brazil, on 6-January-2021 (1). P.1 was eventually reported in Manaus city (Amazonas state capital), on 11-January-2021 (2). Later, it was identified in samples collected since 6-Dec-2020 from Manaus (3). According to phylogenetic studies, P.1 likely emerged in the Amazonas state in early (3) or late (4) November 2020. This variant shares mutations with other variants of concern (VOCs) previously detected in the United Kingdom and South Africa (B.1.1.7 and B.1.351, respectively) (2). Mutations of these two other variants are associated with greater transmissibility and immune evasion (5, 6), which confer them the status of variant of concern. However information, data, and analyzes on the epidemiology of P.1 are still incipient.

The Coronavirus disease 2019 (COVID-19) outbreak in Manaus (April–May 2020) was followed by a period of high but stable incidence, after which prevalence may have reached 42% (7) to 76% (8) by November 2020. From December 2020 to February 2021 the city was devastated by a new outbreak that caused a collapse in the already fragile health system (9), with shortages of oxygen supply (10), while the frequency of P.1 increased sharply from 0% in November 2020 to 73% in January 2021 (4). The pathogenicity of P.1 variant is still unknown, although recent studies point to increased viral load in individuals infected with the new variant (4), suggesting it could be higher than the one from previous circulating strain. We analyzed Brazilian national health surveillance data on COVID-19 hospitalizations and the frequency of P.1 among sequences from residents of Manaus city using a model-based approach (an extended SEIR compartmental model – see Fig. 1) to estimate the transmissibility and relative force of reinfection of the P.1 variant.

**Fig. 1.**
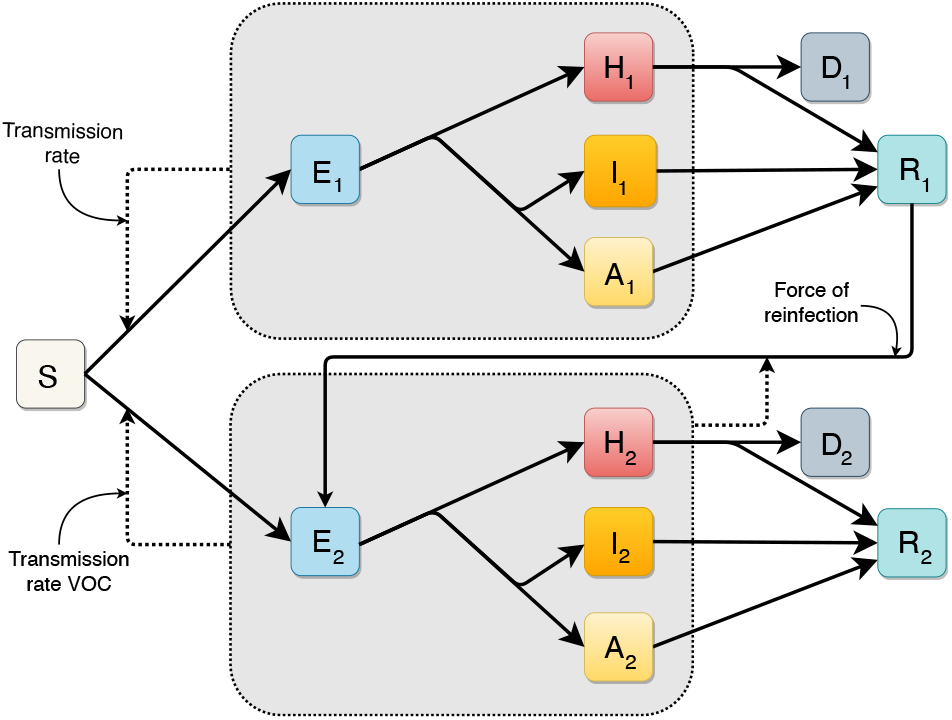
Diagram of the extended deterministic compartmental model (SEAIHRD). S: Susceptible, E: Exposed (pre-symptomatic), H: Hospitalized (severe infected individuals), I: Infected (symptomatic individuals, not hospitalized), A: Asymptomatic. D: Deceased, R: Recovered. Compartments are subdivided into 3 age categories, not represented here for simplicity. Compartments with subindex 1 represent the wild-type variant, subindex 2 refers to the VOC P.1. Continuous lines represent flux between each compartment; dashed lines, infection pathways. Small arrows indicate force of reinfection and transmissibility.

**Fig. 2.**
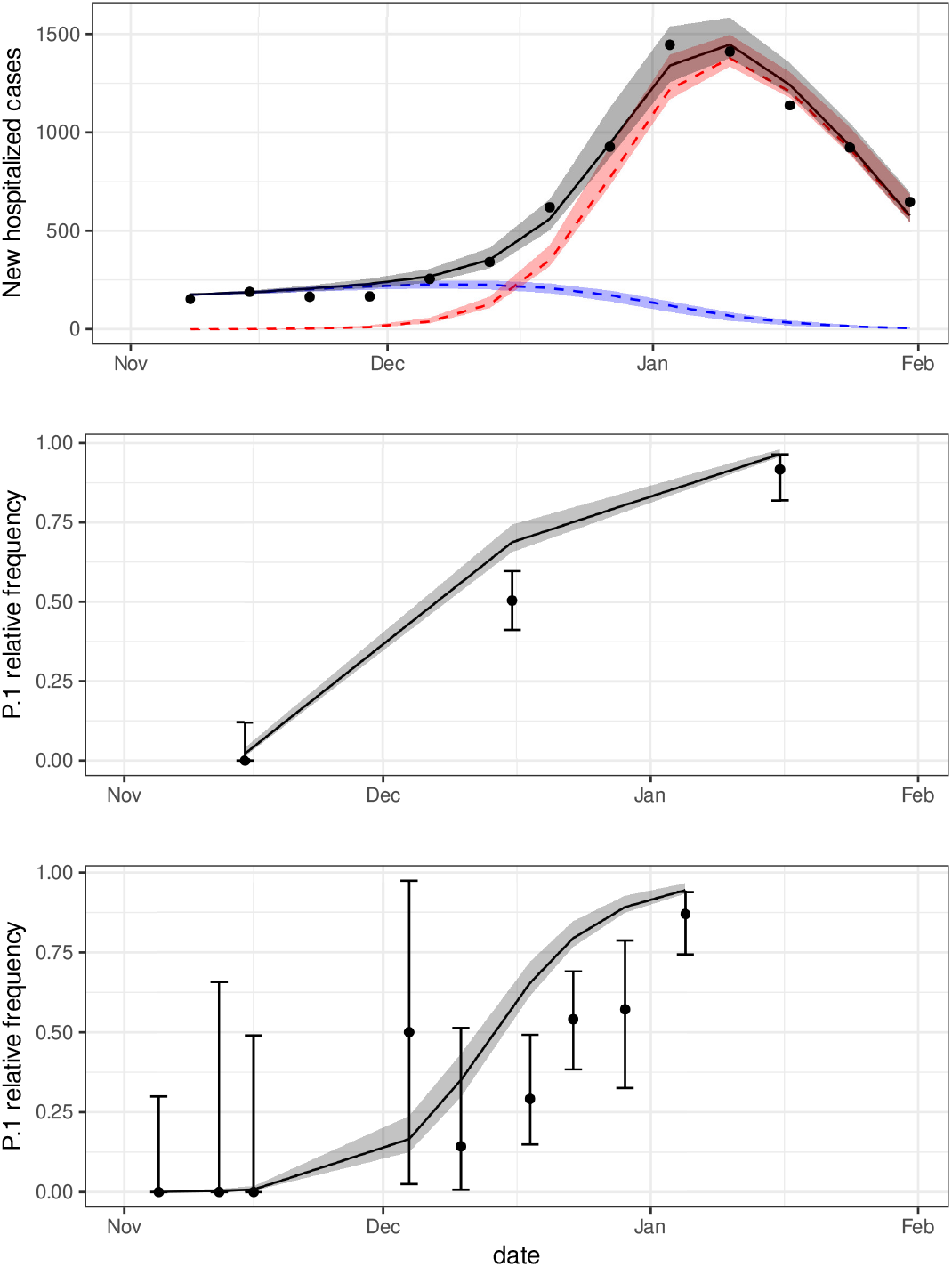
First panel: Weekly new hospitalized COVID-19 cases in Manaus city. Grey line represents the fitted values of total cases (all variants) by maximum likelihood estimation (MLE) of the parameters. Red and blue lines represent the predicted values of cases due to P.1 and wild-type variants, respectively. Black dots are nowcasted observed data of hospitalizations. Second and third panels show the fittings to the time-series frequency of P.1 on datasets provided by (11) and (3) respectively. The area around the lines indicate the 95% Confidence Interval (CI) of the expected values. Dots and lines are the sample proportions of P.1 in sequenced genomes, and their 95% sample CI. The fitted values of the model parameters are presented in Table 1.

## Results

The estimated transmissibility of P.1 was 2.6 (95% Confidence Interval (CI): 2.4—2.8) times higher compared to the wild-type variant, while the relative force of reinfection of the new variant was estimated to be 0.032 (CI: 0.026— 0.040, Table 1) The fitted model also estimated that, at the time P.1 variant emerged, the prevalence of previous infection by the wild-type variant was 78% (CI: 73–83%), and that the number of cases by the wild-type variant were increasing with an estimated daily intrinsic growth rate of 0.029 days^−1^ (CI: 0.024—0.035 days^−1^). Given these parameter values, reinfections by P.1 accounted for 28% of the cases in Manaus from November 2020 through January 2021.

**Table 1.** **Summary of the fitted parameters and respective confidence intervals considering the entire period, November-1, 2020–January-31, 2021 maintaining the same pathogenicity of the previous variant. Sensitivity analyses were performed considering different pathogenicity of the P.1 variant (SA1) and data censuring after the collapse of the healthcare system (SA2) in Manaus, Brazil, on 10-Jan-2021.** **\* parameter was fixed, not estimated, in this analysis**

| Parameter | main fitting |  |  | SA1 |  |  | SA2 |  |  |
| --- | --- | --- | --- | --- | --- | --- | --- | --- | --- |
|  | estimate | 2.5 % | 97.5% | estimate | 2.5 % | 97.5 % | estimate | 2.5 % | 97.5 % |
| Relative transmission rate for the new variant | 2.61 | 2.45 | 2.76 | 2.52 | 2.28 | 2.76 | 2.95 | 2.70 | 3.20 |
| Relative force of reinfection of P.1 | 0.032 | 0.026 | 0.040 | 0.053 | 0.044 | 0.065 | 0.000 | 0.000 | 0.000 |
| Prevalence of previous infection (2020-11-01) (%) | 78 | 73 | 83 | 73 | 67 | 78 | 71 | 69 | 74 |
| Initial fraction of the new variant (2020-11-01) ($\times 10^{-5}$) | 30.4 | 8.2 | 112.9 | 8.5 | 1.4 | 50.8 | 17.6 | 5.0 | 62.4 |
| Intrinsic growth rate (days$^{-1}$) | 0.029 | 0.024 | 0.035 | 0.045 | 0.037 | 0.052 | 0.030 | 0.026 | 0.034 |
| Relative IHR odds ratio | 1* | – | – | 0.74 | 0.63 | 0.85 | 1* | – | – |

We also evaluated the impact of a distinct pathogenicity of the P.1 variant on our estimates by allowing the infection hospitalization rate (IHR) of the new variant to be estimated as a free parameter (see SA1 in Table 1). The relative transmissibility and prevalence did not differ statistically from the the previous estimates. However, the data gives no support for a higher IHR of P.1. Moreover, the model fit to hospitalization data prior to the healthcare system collapse in the city of Manaus (11 January, 2021) estimated an even larger transmissibility (SA2 in Table 1).

## Discussion

COVID-19 hospitalizations and frequency of the P.1 variant in clinical samples showed a sharp increase in Manaus, Brazil, starting December 2020. The fitted model describes this joint increase as the result of the emergence of P.1, estimated to be 2.6 times more transmissible than the wild-type variant. The spread of P.1 occurred despite a high estimated prevalence of infection by the wild-type virus. The pathogenicity of P.1 is still unknown, but assuming hospitalization rates as a proxy for pathogenicity, P.1 transmissibility holds for different ranges of pathogenicity. Two recent studies analysed genomic data of SARS-CoV-2 from Manaus evaluating the transmissibility of the new variant (3, 4). Faria and collaborators integrated mortality and genomic data and, using a semi-mechanistic Bayesian model, estimated a transmissibility 1.4–2.2 times higher and 25–61% evasion of protective immunity related to the variant P.1 (3). Naveca and collaborators estimated a 2.2 times higher effective reproduction number for the P.1 variant using phylogenetic methods, and suggested that P.1 is at least two times more transmissible than the parental lineage, assuming reinfections are rare (4). The present work follows a different approach that can be defined as an epidemiological, model-based, and data-fitting approach, suitable for scenarios where only surveillance data are available, and applicable to other emerging variants throughout the world. Notably, all three different approaches estimated very high transmissibility of the P.1 variant.

Many knowledge gaps about the pandemic in the Amazonian region still remain. Population-based serological surveys are not available and thus prevalence was included in the fitted parameters. The analysed data overlapped with the period of the health system collapse. Aware that in-hospital fatality rates can quickly change when the health system is under stress (12), we have chosen hospitalization data instead of mortality data (See **Dataset**). Still, during the health system collapse many severe cases probably were not recorded in the system and remained unaccounted for. Our results were robust to removing this period in the sensitivity analysis (SA2). Even without P.1 emergence, the model estimates an increase in the number of cases (parameter *r*, see Table 1), which could be a consequence of loosening non-pharmacological interventions (NPIs) (9), an effect of waning immunity, or both. Our model does not consider these effects explicitly, but by fitting the initial growth rate we indirectly account for their effects on the dynamics and on the estimation of the remaining parameters.

The impacts of a highly transmissible variant have already been highlighted by the spread of VOC B.1.1.7 in the UK, USA and Europe (13). The variant B.1.1.7 has an upper-bound estimate for transmissibility of 2.3 (5), which is smaller than our lower bound estimate for P.1. Higher transmissibility of the P.1 variant raises strong concerns of swift upsurges in the number of cases once P.1 reaches community transmission. Although our estimate for the relative force of reinfection by the variant P.1 seems low, the impact is strong enough to drive, together with a high transmissibility, a large surge even in a population heavily affected by the wild-type variant. For instance, in Manaus, 28% of the new cases in the period considered were due to reinfections by P.1 in our estimations, reaching 40% when assuming a different IHR for P.1 (SA1). However, in a scenario of low prevalence rate of infection by the wild-type variant, the high transmissibility is the most determinant parameter of the rapid increase in the number of cases and can lead to even sharper outbreaks. The P.1 variant has already been detected in at least 26 countries, with local transmission currently confirmed in four of them (13). This points to the urgency of reinforcing measures to avoid a global spread of P.1, which include an agile global genomic surveillance network. Further, to improve our ability to deal with the threat of P.1, it is urgent to study i) the pathogenicity of the P.1 variant, since this trait, in association with high transmissibility, can drive even well-prepared health systems to collapse; ii) the efficacy of current vaccines for P.1 variant infections; and iii) the main factors promoting the emergence of VOCs, specially the roles of previous high prevalence and of waning immunity.

## Materials and Methods

### Dataset

We used the Brazilian epidemiological syndrome surveillance system for influenza, SIVEP-Gripe (https://opendatasus.saude.gov.br), to track COVID-19 hospitalized cases. All hospitalized patients with Severe Acute Respiratory Illness (SARI) are reported to SIVEP-Gripe with symptom onset date and SARS-CoV-2 test results. SIVEP-Gripe, due to its universal coverage and mandatory notification of SARI cases, has an homogeneous testing effort to diagnose SARS-CoV-2 infections, and is currently the best source for Brazilian data at the national level. Hospitalization data provides the most accurate basis to infer incidence in Manaus, because mild cases are vastly under-reported and testing capacity fluctuates, while mortality data is harder to relate to total number of cases, since the city’s health system endured a prolonged stress even before the collapse, with large variation in the in-hospital fatality rate over time (12). Data for hospitalized COVID-19 cases among residents in Manaus from 01-Nov-2020 to 31-Jan-2021 was obtained from SIVEP-Gripe database of 15-Feb-2021. The hospitalized cases of the last 10 weeks in the time series were nowcasted (14) to correct for notification delay. Time-series of frequency of sequenced genomes identified as P.1 in Manaus were extracted from published datasets (3, 11).

### Model

A deterministic compartmental model (Figure 1) was developed to model the infection of Susceptible individuals moving to the Exposed (pre-symptomatic) compartment, which can progress to three alternative compartments: Hospitalized (severely ill), Infected (symptomatic but non-hospitalized), and Asymptomatic. Eventually, individuals move to Recovered or Deceased. Two variants are considered: 1-non-P.1 (“wild-type”) and 2-new/P.1. The latter is assumed to infect Recovered individuals previously infected by the wild-type, and no reinfections of wild-type due to waning immunity occur. Compartments were stratified into three age categories: young (< 20 years old), adult (≥ 20 and < 60 years old) and elderly (≥ 60 years old), with different rates for outcomes. The key parameters of relative transmissibility and relative force of reinfection – the ratio between the force of infection by P.1 on previously infected individuals (reinfections) and the force of infection by P.1 on susceptible ones (new infections) – were estimated by a maximum likelihood fitting to the weekly number of new hospitalizations and to genomic surveillance data. Three additional model parameters with unknown values were estimated. The remaining parameters (24 out of 29) were fixed, using current values from the literature (see Supplementary Information (SI) for values and references). Sensitivity to different pathogenicity of the variant P.1 was explored by repeating the fit assuming IHR as a free parameter (SA1). The sensitivity to the period analysed was also explored by another fit excluding the health system collapse period (SA2). Further model details and fitting methodology are available in the SI.

## Data Availability

All data used are publicly available. The data sources are described in the manuscript and in supplementary file.

## ACKNOWLEDGMENTS

We are grateful for the collaborative work of the entire group of the Observatório COVID-19 BR. In particular, we thank Verônica Coelho for critical inputs. The authors also thank the research funding agencies: the Coordenação de Aperfeiçoamento de Pessoal de Nível Superior – Brazil (Finance Code 001 to FMDM, LSF and TPP), Conselho Nacional de Desenvolvimento Científico e Tecnológico – Brazil (grant number: 315854/2020-0 to MEB, 141698/2018-7 to RLPS, 313055/2020-3 to PIP, 312559/2020-8 to MASMV, 311832/2017-2 to RAK, 305703/2019-6 to AAMS) and Fundação de Amparo à Pesquisa do Estado de São Paulo - Brazil (grant number: 2019/26310-2 and 2017/26770-8 to CF, 2018/26512-1 to OC, 2018/24037-4 to SP and contract number: 2016/01343-7 to RAK). **Disclaimer** The findings and conclusions in this article are those of the authors and do not necessarily represent the official position of the Centers of Disease Control and Prevention.

## Supplementary Information for

## Supporting Information Text

In order to estimate key parameters of the variant of concern (VOC) P.1, we developed a model and fitted it to time-series data of number of hospitalized cases and frequency of the P.1 variation. The fitting approach used here can be applied to other regions where data is relatively scarce. It primarily requires weekly incidence data to determine proper model initial conditions. In Brazil, these are the hospitalized cases data. Stratification by age allows the model to also consider the different death rates, asymptomatic and hospitalized proportions of each age class, important features for SARS-CoV-2. Contact levels between different age classes, which may vary from one place to another, can also be considered. For special cases in which information such as contact between ages classes and age distribution are not available (or even unnecessary for some other disease), the model can be easily simplified. In this sense, the method proposed here demands low-detailed data and relies on the structure of a simple compartmental model to measure quantities of interest, such as transmissibility and relative force of reinfection.

Section 1 describes the model, section 2 relates the values of the parameters taken from the current literature, section 3 describes the contact matrix used, and finally section 4 describes the treatment of case data (subsec. A), the choice of initial conditions (subsec. B), the fitting procedure (subsec. C), and the sensitivity analysis evaluated regarding pathogenicity and data period analysed (subsec. D)

## 1. Model equations

The model is an extended Susceptible, Exposed, Infected, and Recovered (SEIR) model that comprises susceptible (S), pre-symptomatic (E), asymptomatic (A), mild symptomatic (I), severe/hospitalized (H), recovered (R) and deceased (D) compartments. These compartments are duplicated to account for a second variant of SARS-CoV-2, and each of them is stratified into three age classes: young (<20 years old), adults ([20 *—* 59] years old), and the elderly (≥ 60 years old). The “wild-type” classes represent all non-P.1 variants present, which do not seem to be variants of concern.

We assume that the second variant is capable of reinfecting individuals who have recovered from infection by the wild-type variant while the inverse is not possible; in the absence of data indicating this possibility, allowing reinfection by the wild-type variant on recovered of infection by P.1 would have negligible effect due to the small time window (3 months) considered in the present work. We also consider that a variant is not capable of reinfecting individuals recovered from the same lineage. Our model does not include vaccination due to low rates of vaccination in Brazil during the study time period.

To model the virus spread in the population, we assume that asymptomatic individuals have equal infectiousness compared to symptomatic ones, while pre-symptomatic individuals have reduced infectiousness represented by *ω*. To model behaviour, we assume that symptomatic individuals self-isolate themselves to some degree, reducing their contacts by *ξ*. Individuals with severe disease have greater isolation *ξ*_*sev*_ due to hospitalization. The daily contacts between each age class is represented by the matrix *Ĉ*. The force of infection *λ*_*k*_ for each variant *k* is defined below:

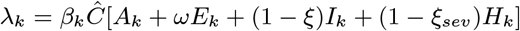

The complete system of equations is given by:

### Completely Susceptible

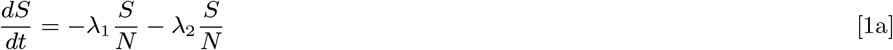

### Wild variant

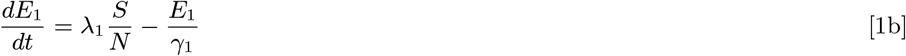

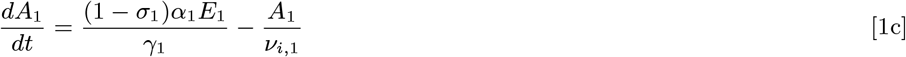

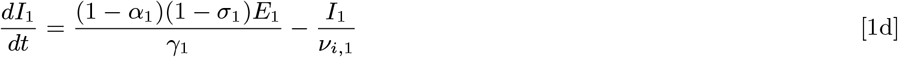

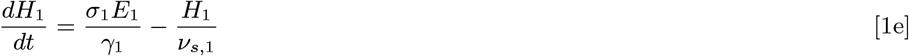

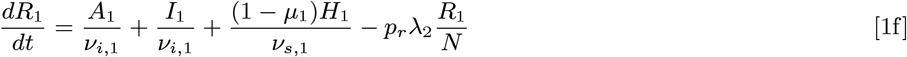

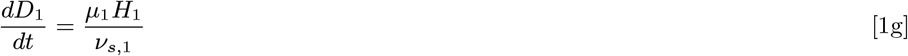

### P.1 variant

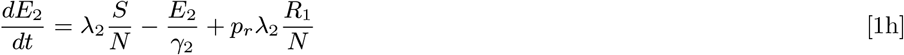

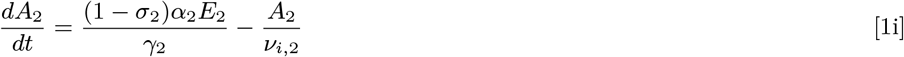

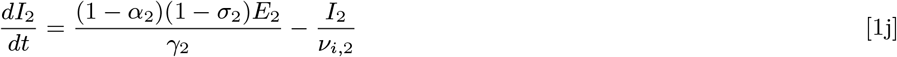

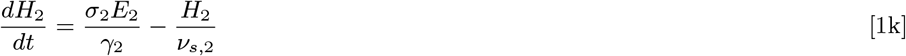

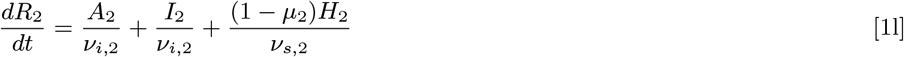

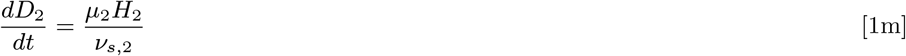

### Supplementary Equations

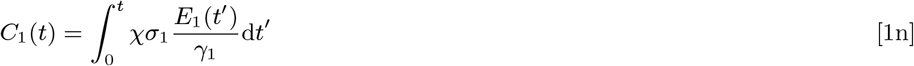

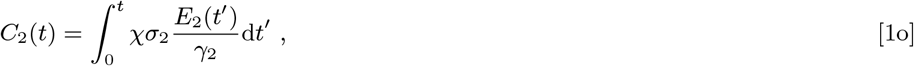

where *C*_1_ ad *C*_2_ are the cumulative hospitalization cases reported, and each variable of the system (*S, E*_*k*_, …, *C*_*k*_) is a vector containing each age class, *e*.*g*., *E*_1_ = (*E*_1,*y*_, *E*_1,*a*_, *E*_1,*e*_)^*T*^. The equations were numerically solved by the R package developed by (1).

## 2. Parameterization of the model

The parameters considered for the wild-variant are described below. The parameters for the P.1 variant are the same except for those considered in the model fitting.

- *γ*, Average time in days between being infected and developing symptoms: 5.8 (2)
- *ν*_*i*_, Average time in days between being infectious and recovering for asymptomatic and mild cases: 9.0 (3)
- *ν*_*s*_, Average time between being infectious and recovering/dying for severe cases: 8.4 SIVEP-Gripe for São Paulo State
- *ξ*, reduction on the exposure of symptomatic cases (due to symptoms/quarantining): 0.1 [Assumed]
- *ξ*_*sev*_, Reduction on the exposure of severe cases (due to hospitalization): 0.9 [Assumed]
- *ω*, Relative infectiousness of pre-symptomatic individuals: 1.0 [Assumed]
- *α*, Proportion of asymptomatic cases [0.67,0.44,0.31] for Juvenile (4), Adult and Elderly (5)
- *σ*, Proportion of infections that require hospitalization: [0.001,0.012,0.089]* (6)
- *µ*, In-hospital mortality ratio: [0.417,0.188,0.754] (7)
- *χ*, Case report probability: 1.0 [Assumed]

^*\**^The proportion is weighted by the age distribution of the population with each age category.

### 3. Contact Matrices

Our model includes three age group categories: young ([0 *—* 19] *y*.*o*.), adults ([20 *—* 59] *y*.*o*.), and elderly (greater than 60*y*.*o*.). To model contacts between these groups we use estimated contact matrices computed by (8), but since the original matrices use five-year age bins going up to 95+ years, we aggregate classes leading to a 3 *×* 3 matrix in the following way:

Let *A, B* be sets of indexes forming age groups (not necessarily of equal sizes), *x*_*i,j*_ denoting contact between age groups *i* and *j* in the original matrix, *d*_*i*_ denoting population size of the age group *i*. The new contact matrix *Ĉ* is given by:

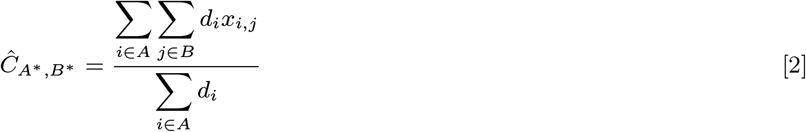

where *A*^*^, *B*^*^ denotes a new indexation rule. Note that the contact matrices depend on local demographics and therefore must be computed for each place of study.

## 4. Data Analysis Procedures

### A. Nowcasting

Data used in parameters estimation were collected from the national public health system of severe acute respiratory illness (SARI) surveillance database, named *Sistema de Vigilância da Gripe - SIVEP-Gripe*. In this system, reporting of cases can be delayed for several reasons, including the notification system itself and confirmation of RT-PCR test results. The nowcasting procedure estimates, based on the past delay distribution, the number of cases that already occurred but were not yet reported. A window of 10 weeks is the acting window on the series, since delays greater than this are rare.

Nowcasting requires a pair of dates: (i) onset date of the event and (ii) report date of the event. The delay distribution is modeled as being best described as a Poisson distribution for days since the onset date to the report date. We considered *the first symptoms date* as the onset date. For the report date, we used the latest between *the test result date* and *the clinical classification date*. The nowcasting algorithm were developed by (9), and implemented in the NobBS (Nowcasting by Bayesian Smoothing) package in R (10).

### B. Initial Condition Estimation

The model requires appropriate mid-epidemic initial conditions in order to give relevant results. In the model, the number of new hospitalizations at a given time – *h*_*new*_, is directly proportional to the number of exposed individuals at that time, therefore data was used to get an approximation of the number of exposed people. Also, to quantify the number of people belonging to the recovered class, prevalence was used.

We can estimate the appropriate initial conditions by finding an approximation for our model that relates more directly to the available data in each class. In the absence of the variant P.1, the model has four classes of infected compartments, namely **y** = (*E*_1_, *A*_1_, *I*_1_, *H*_1_)^*T*^, and another three classes, represented by **z**, i.e., **z** = (*S, R*_1_, *D*_1_)^*T*^. To that effect, we can write the system as

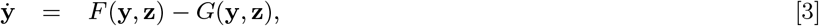

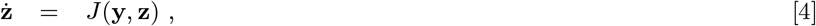

where *F* comprises all entries of new Infected, coming from classes **z**, whilst *G* accounts for the transitions within infected classes and also recovery and death from the disease. Then, to find a good approximation for a small time window, we perform a linearization of our model around a point (**y, z**). Keeping **z** fixed, we get

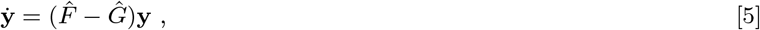

where 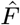 and *Ĝ* are the linearized matrices arising from the functions *F* and *G*, respectively. The only entrance of new infected comes from the *βSλ/N* terms in the *Ė*_1_ = (*Ė*_1,*y*_, *Ė*_1,*a*_, *Ė*_1,*e*_)^*T*^ equations (sub-indexes are *y* young, *a* adults and *e* elderly), then, the only non-zero elements of 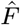 are in its first 3 lines. Before proceeding, it is useful to define

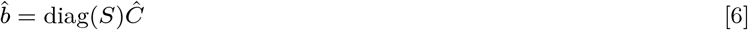

which allow us to write

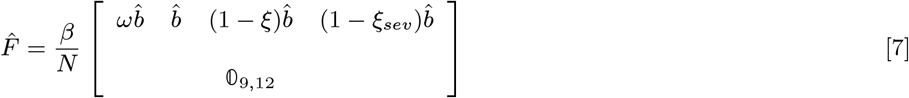

*Ĝ* contains the terms of Exposed, *E*_1_, the 3 possible forms of the disease considered in the model (*A*_1_, *I*_1_ and *H*_1_), as the terms in its first 3 rows,, whilst the remainder of its main diagonal contains terms of recovery and death. For simplicity, every constant (or vector for the terms with *σ*) in *Ĝ* expression Eq. (8) should be thought as diagonal matrices with its elements given by the constants (or vectors) and every *0* is a 3-dimensional square matrix where all entries are null.

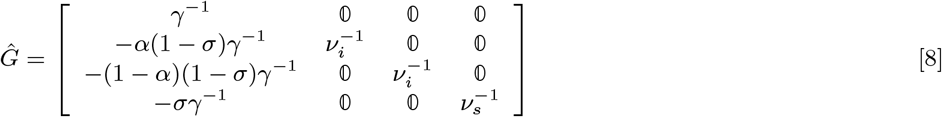

The linearization above implies that, for a small time interval, **y** has an exponential behavior and that the eigenvalues of 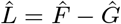 are related to the exponential growth rates. Therefore, a short time after the beginning of the epidemic, the largest eigenvalue should be the one to dominate. So the exponential growth rate of the wild-type variant – *r*, can be matched to the largest eigenvalue of 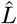 to obtain an estimate for *β*. The eigenvector associated with the largest eigenvalue gives the proportions of infected classes, which, together with the estimated number of exposed individuals – *E*_1_ = *γ*_1_*h*_*new*_ */σ*_1_, results in an approximation for the number of people in the other infected classes.

Given a *β*, the largest eigenvalue of the linearization matrix is computed using the <monospace>eigs </monospace>function of the R package rARPACK (11) and we find the *β* that gives *r* as the largest eigenvalue through bisection root finding. Finally, subtracting the number of recovered and infected from the total population gives the number of susceptible individuals.

### C. Maximum Likelihood Estimation

Given the cumulative daily curves of hospitalization for wild-type variant, *C*_1_, and P.1 variant, *C*_2_, we can obtain the daily variation of each curve, namely 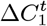 and 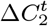. Those curves are summed up to give the total number of weekly new cases:

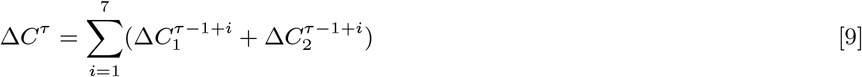

where *τ* is a discrete index given in weeks.

To calculate the frequency of P.1 in a given time period *T*, we use the proportion of new cases in this period from the wild-type and P.1 variant as follows:

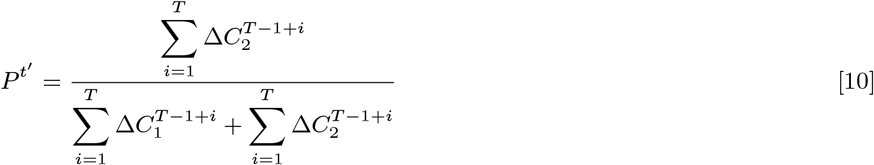

where *t*^I^ is a discrete index given in *T* periods.

The time period *T* depends on the dataset of genome sequences: it is daily in (12) and monthly in (13).

Using maximum likelihood, we fitted the model by estimating five parameters, namely, the relative transmissibility 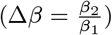, the relative force of reinfection of P.1 (*p*_*r*_), initial total prevalence (*ρ*^0^ = [*R/N*]_*t*=0_), initial fraction of cases that were caused by the new variant (*P* ^0^), and intrinsic growth rate of the wild-type variant (*r*). The initial fraction of P.1 cases (*P* ^0^) accounts for the uncertainty in the time of emergence of the new variant: the simulation starts at beginning of November, so this initial value is below 1 individual, and only reaches this threshold by mid-to late November, depending on the value of *P* ^0^. The parameter *r* incorporates effects related to contact rates for the wild-type variant, such as non-pharmacological interventions relaxation, elections, and others; it affects the transmissibilities of both variants (*β*_1_ and *β*_2_) in the same way, and so is independent of Δ*β*.

Number of hospitalization cases were assumed to follow a Poisson distribution, with expected value given by equation Eq. (9). The recorded number of P.1 in genome samples was assumed to follow a binomial distribution with an expected value equal to the product of the total number of genome sequences sampled in each date and the proportion of P.1 cases (equation Eq. (10)). The log-likelihood function for the model fitting was then:

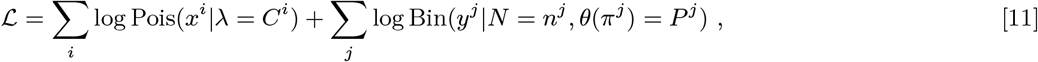

where Pois is a Poisson distribution with parameter *λ, x*^*i*^ is the number of recorded hospitalizations in week *i*, Bin is a Binomial distribution with parameters *N* (total number of trials) and *π*^*j*^ (probability of success at each trial), *n*^*j*^ is total number of sequences in clinical samples in week or day *j, y*^*j*^ is the number of P.1 sequences in each of these samples, and *θ*(.) is the logit function.

The model was then fitted by finding the values of the five above mentioned parameters that minimize the negative of the log-likelihood function (equation 11), using the function mle2, from the R package bbmle (14).

To find starting values for the optimization performed by mle2 we calculated the log-likelihood function for one million combinations of parameters values in a regular reticulate within reasonable ranges. The 100 sets of parameters that were local minima and with highest log-likelihood were used as starting values for the computational minimization.

The confidence intervals for the expected number of cases and frequency were estimated from 20000 parametric bootstrap samples assuming that the estimated parameters follow a multivariate normal distribution. The parameters of these multivariate distributions were the estimated values and estimated variance-covariance matrix of the parameters. For each sampled combination of para 2.5% and 95% quantiles of the distribution of bootstrapped expected values.

### D. Sensitivity analysis

The model fitting assumed a constant infection hospitalization rate (IHR, parameter *σ*) for each age group over time for both variants. Recent evidence suggests that prior SARS-CoV-2 infection protects most individuals against reinfection (15), so reinfections might have lower IHR. Because the pathogenicity of the variant P.1 is unknown, the model fitting was repeated assuming that the odds ratio of the IHR in each age class for P.1 infections compared to wild-type variant infections (SA1) is a free parameter. Moreover, as the collapse of Manaus health system hindered hospitalizations of new severe cases and may have affected case recording in surveillance databases, the model fitting was repeated considering only the period prior to the collapse (10-January-2021) (SA2).

