## Supplementary file for "Model-based estimation of transmissibility and reinfection of SARS-CoV-2 P.1 variant"

To model the virus spread in the population, we assume that asymptomatic individuals have equal infectiousness compared to symptomatic ones, while pre-symptomatic individuals have reduced infectiousness represented by $\omega$. To model behaviour, we assume that symptomatic individuals self-isolate themselves to some degree, reducing their contacts by $\xi$. Individuals with severe disease have greater isolation $\xi_{sev}$ due to hospitalization. The daily contacts between each age class is represented by the matrix $\hat{C}$. The force of infection $\lambda_k$ for each variant $k$ is defined below:

$$\lambda_k = \beta_k \hat{C}[A_k + \omega E_k + (1-\xi)I_k + (1-\xi_{sev})H_k]$$

The complete system of equations is given by:

**Completely Susceptible**

$$\frac{dS}{dt} = -\lambda_1 \frac{S}{N} - \lambda_2 \frac{S}{N} \quad [1a]$$

**Wild variant**

$$\frac{dE_1}{dt} = \lambda_1 \frac{S}{N} - \frac{E_1}{\gamma_1} \quad [1b]$$

$$\frac{dA_1}{dt} = \frac{(1-\sigma_1)\alpha_1 E_1}{\gamma_1} - \frac{A_1}{\nu_{i,1}} \quad [1c]$$

$$\frac{dI_1}{dt} = \frac{(1-\alpha_1)(1-\sigma_1)E_1}{\gamma_1} - \frac{I_1}{\nu_{i,1}} \quad [1d]$$

$$\frac{dH_1}{dt} = \frac{\sigma_1 E_1}{\gamma_1} - \frac{H_1}{\nu_{s,1}} \quad [1e]$$

$$\frac{dR_1}{dt} = \frac{A_1}{\nu_{i,1}} + \frac{I_1}{\nu_{i,1}} + \frac{(1-\mu_1)H_1}{\nu_{s,1}} - p_r \lambda_2 \frac{R_1}{N} \quad [1f]$$

$$\frac{dD_1}{dt} = \frac{\mu_1 H_1}{\nu_{s,1}} \quad [1g]$$

**P.1 variant**

$$\frac{dE_2}{dt} = \lambda_2 \frac{S}{N} - \frac{E_2}{\gamma_2} + p_r \lambda_2 \frac{R_1}{N} \quad [1h]$$

$$\frac{dA_2}{dt} = \frac{(1-\sigma_2)\alpha_2 E_2}{\gamma_2} - \frac{A_2}{\nu_{i,2}} \quad [1i]$$

$$\frac{dI_2}{dt} = \frac{(1-\alpha_2)(1-\sigma_2)E_2}{\gamma_2} - \frac{I_2}{\nu_{i,2}} \quad [1j]$$

$$\frac{dH_2}{dt} = \frac{\sigma_2 E_2}{\gamma_2} - \frac{H_2}{\nu_{s,2}} \quad [1k]$$

$$\frac{dR_2}{dt} = \frac{A_2}{\nu_{i,2}} + \frac{I_2}{\nu_{i,2}} + \frac{(1-\mu_2)H_2}{\nu_{s,2}} \quad [1l]$$

$$\frac{dD_2}{dt} = \frac{\mu_2 H_2}{\nu_{s,2}} \quad [1m]$$

**Supplementary Equations**

$$C_1(t) = \int_0^t \chi\sigma_1 \frac{E_1(t')}{\gamma_1} \mathrm{d}t' \quad [1n]$$

$$C_2(t) = \int_0^t \chi\sigma_2 \frac{E_2(t')}{\gamma_2} \mathrm{d}t' \ , \quad [1o]$$

where $C_1$ ad $C_2$ are the cumulative hospitalization cases reported, and each variable of the system ($S$, $E_k, ...,$ $C_k$) is a vector containing each age class, *e.g.*, $E_1 = (E_{1,y}, E_{1,a}, E_{1,e})^T$. The equations were numerically solved by the R package developed by (1).

### 2. Parameterization of the model

The parameters considered for the wild-variant are described below. The parameters for the P.1 variant are the same except for those considered in the model fitting.

- $\gamma$, Average time in days between being infected and developing symptoms: 5.8 (2)
- $\nu_i$, Average time in days between being infectious and recovering for asymptomatic and mild cases: 9.0 (3)
- $\nu_s$, Average time between being infectious and recovering/dying for severe cases: 8.4 SIVEP-Gripe for São Paulo State
- $\xi$, reduction on the exposure of symptomatic cases (due to symptoms/quarantining): 0.1 [Assumed]
- $\xi_{sev}$, Reduction on the exposure of severe cases (due to hospitalization): 0.9 [Assumed]
- $\omega$, Relative infectiousness of pre-symptomatic individuals: 1.0 [Assumed]
- $\alpha$, Proportion of asymptomatic cases [0.67,0.44,0.31] for Juvenile (4), Adult and Elderly (5)
- $\sigma$, Proportion of infections that require hospitalization: [0.001,0.012,0.089]* (6)
- $\mu$, In-hospital mortality ratio: [0.417,0.188,0.754] (7)
- $\chi$, Case report probability: 1.0 [Assumed]

*The proportion is weighted by the age distribution of the population with each age category.

### 3. Contact Matrices

Our model includes three age group categories: young ($[0-19]\, y.o.$), adults ($[20-59]\, y.o.$), and elderly (greater than $60 y.o.$). To model contacts between these groups we use estimated contact matrices computed by (8), but since the original matrices use five-year age bins going up to 95+ years, we aggregate classes leading to a $3 \times 3$ matrix in the following way:

Let $A$, $B$ be sets of indexes forming age groups (not necessarily of equal sizes), $x_{i,j}$ denoting contact between age groups $i$ and $j$ in the original matrix, $d_i$ denoting population size of the age group $i$. The new contact matrix $\hat{C}$ is given by:

$$\hat{C}_{A^*,B^*} = \frac{\sum_{i\in A}\sum_{j\in B} d_i x_{i,j}}{\sum_{i\in A} d_i} \quad [2]$$

where $A^*, B^*$ denotes a new indexation rule. Note that the contact matrices depend on local demographics and therefore must be computed for each place of study.

### 4. Data Analysis Procedures

**A. Nowcasting.** Data used in parameters estimation were collected from the national public health system of severe acute respiratory illness (SARI) surveillance database, named *Sistema de Vigilância da Gripe - SIVEP-Gripe*. In this system, reporting of cases can be delayed for several reasons, including the notification system itself and confirmation of RT-PCR test results. The nowcasting procedure estimates, based on the past delay distribution, the number of cases that already occurred but were not yet reported. A window of 10 weeks is the acting window on the series, since delays greater than this are rare.

Nowcasting requires a pair of dates: (i) onset date of the event and (ii) report date of the event. The delay distribution is modeled as being best described as a Poisson distribution for days since the onset date to the report date. We considered *the first symptoms date* as the onset date. For the report date, we used the latest between *the test result date* and *the clinical classification date*. The nowcasting algorithm were developed by (9), and implemented in the NobBS (Nowcasting by Bayesian Smoothing) package in R (10).

**B. Initial Condition Estimation.** The model requires appropriate mid-epidemic initial conditions in order to give relevant results. In the model, the number of new hospitalizations at a given time – $h_{new}$, is directly proportional to the number of exposed individuals at that time, therefore data was used to get an approximation of the number of exposed people. Also, to quantify the number of people belonging to the recovered class, prevalence was used.

We can estimate the appropriate initial conditions by finding an approximation for our model that relates more directly to the available data in each class. In the absence of the variant P.1, the model has four classes of infected compartments, namely $\mathbf{y} = (E_1,\ A_1,\ I_1,\ H_1)^T$, and another three classes, represented by $\mathbf{z}$, i.e., $\mathbf{z} = (S,\ R_1,\ D_1)^T$. To that effect, we can write the system as

$$\dot{\mathbf{y}} = F(\mathbf{y},\mathbf{z}) - G(\mathbf{y},\mathbf{z}), \qquad [3]$$

$$\dot{\mathbf{z}} = J(\mathbf{y},\mathbf{z})\ , \qquad [4]$$

where $F$ comprises all entries of new Infected, coming from classes $\mathbf{z}$, whilst $G$ accounts for the transitions within infected classes and also recovery and death from the disease. Then, to find a good approximation for a small time window, we perform a linearization of our model around a point $(\mathbf{y},\mathbf{z})$. Keeping $\mathbf{z}$ fixed, we get

$$\dot{\mathbf{y}} = (\hat{F} - \hat{G})\mathbf{y}\ , \qquad [5]$$

where $\hat{F}$ and $\hat{G}$ are the linearized matrices arising from the functions $F$ and $G$, respectively. The only entrance of new infected comes from the $\beta S\lambda/N$ terms in the $\dot{E}_1 = (\dot{E}_{1,y}, \dot{E}_{1,a}, \dot{E}_{1,e})^T$ equations (sub-indexes are $y$ young, $a$ adults and $e$ elderly), then, the only non-zero elements of $\hat{F}$ are in its first 3 lines. Before proceeding, it is useful to define

$$\hat{b} = \mathrm{diag}(S)\hat{C} \qquad [6]$$

which allow us to write

$$\hat{F} = \frac{\beta}{N}\begin{bmatrix} \omega\hat{b} & \hat{b} & (1-\xi)\hat{b} & (1-\xi_{sev})\hat{b} \\ & \mathbb{0}_{9,12} & & \end{bmatrix} \qquad [7]$$

$\hat{G}$ contains the terms of Exposed, $E_1$, the 3 possible forms of the disease considered in the model ($A_1, I_1$ and $H_1$), as the terms in its first 3 rows, , whilst the remainder of its main diagonal contains terms of recovery and death. For simplicity, every constant (or vector for the terms with $\sigma$) in $\hat{G}$ expression Eq. (8) should be thought as diagonal matrices with its elements given by the constants (or vectors) and every $\mathbb{0}$ is a 3-dimensional square matrix where all entries are null.

$$\hat{G} = \begin{bmatrix} \gamma^{-1} & \mathbb{0} & \mathbb{0} & \mathbb{0} \\ -\alpha(1-\sigma)\gamma^{-1} & \nu_i^{-1} & \mathbb{0} & \mathbb{0} \\ -(1-\alpha)(1-\sigma)\gamma^{-1} & \mathbb{0} & \nu_i^{-1} & \mathbb{0} \\ -\sigma\gamma^{-1} & \mathbb{0} & \mathbb{0} & \nu_s^{-1} \end{bmatrix} \qquad [8]$$

The linearization above implies that, for a small time interval, $\mathbf{y}$ has an exponential behavior and that the eigenvalues of $\hat{L} = \hat{F} - \hat{G}$ are related to the exponential growth rates. Therefore, a short time after the beginning of the epidemic, the largest eigenvalue should be the one to dominate. So the exponential growth rate of the wild-type variant – $r$, can be matched to the largest eigenvalue of $\hat{L}$ to obtain an estimate for $\beta$. The eigenvector associated with the largest eigenvalue gives the proportions of infected classes, which, together with the estimated number of exposed individuals – $E_1 = \gamma_1 h_{new}/\sigma_1$, results in an approximation for the number of people in the other infected classes.

Given a $\beta$, the largest eigenvalue of the linearization matrix is computed using the `eigs` function of the R package rARPACK (11) and we find the $\beta$ that gives $r$ as the largest eigenvalue through bisection root finding. Finally, subtracting the number of recovered and infected from the total population gives the number of susceptible individuals.

**C. Maximum Likelihood Estimation.** Given the cumulative daily curves of hospitalization for wild-type variant, $C_1$, and P.1 variant, $C_2$, we can obtain the daily variation of each curve, namely $\Delta C_1^t$ and $\Delta C_2^t$. Those curves are summed up to give the total number of weekly new cases:

$$\Delta C^{\tau} = \sum_{i=1}^{7} (\Delta C_1^{\tau-1+i} + \Delta C_2^{\tau-1+i}) \qquad [9]$$

where $\tau$ is a discrete index given in weeks.

To calculate the frequency of P.1 in a given time period $T$, we use the proportion of new cases in this period from the wild-type and P.1 variant as follows:

$$P^{t'} = \frac{\sum_{i=1}^{T} \Delta C_2^{T-1+i}}{\sum_{i=1}^{T} \Delta C_1^{T-1+i} + \sum_{i=1}^{T} \Delta C_2^{T-1+i}} \qquad [10]$$

where $t'$ is a discrete index given in $T$ periods.

The time period $T$ depends on the dataset of genome sequences: it is daily in (12) and monthly in (13).

Using maximum likelihood, we fitted the model by estimating five parameters, namely, the relative transmissibility ($\Delta\beta = \frac{\beta_2}{\beta_1}$), the relative force of reinfection of P.1 ($p_r$), initial total prevalence ($\rho^0 = [R/N]_{t=0}$), initial fraction of cases that were caused by the new variant ($P^0$), and intrinsic growth rate of the wild-type variant ($r$). The initial fraction of P.1 cases ($P^0$) accounts for the uncertainty in the time of emergence of the new variant: the simulation starts at beginning of November, so this initial value is below 1 individual, and only reaches this threshold by mid- to late November, depending on the value of $P^0$. The parameter $r$ incorporates effects related to contact rates for the wild-type variant, such as non-pharmacological interventions relaxation, elections, and others; it affects the transmissibilities of both variants ($\beta_1$ and $\beta_2$) in the same way, and so is independent of $\Delta\beta$.

Number of hospitalization cases were assumed to follow a Poisson distribution, with expected value given by equation Eq. (9). The recorded number of P.1 in genome samples was assumed to follow a binomial distribution with an expected value equal to the product of the total number of genome sequences sampled in each date and the proportion of P.1 cases (equation Eq. (10)). The log-likelihood function for the model fitting was then:

$$\mathcal{L} = \sum_i \log \mathrm{Pois}(x^i|\lambda = C^i) + \sum_j \log \mathrm{Bin}(y^j|N = n^j, \theta(\pi^j) = P^j) \ , \qquad [11]$$

where Pois is a Poisson distribution with parameter $\lambda$, $x^i$ is the number of recorded hospitalizations in week $i$, Bin is a Binomial distribution with parameters $N$ (total number of trials) and $\pi^j$ (probability of success at each trial), $n^j$ is total number of sequences in clinical samples in week or day $j$, $y^j$ is the number of P.1 sequences in each of these samples, and $\theta(.)$ is the logit function.

The model was then fitted by finding the values of the five above mentioned parameters that minimize the negative of the log-likelihood function (equation 11), using the function `mle2`, from the R package bbmle (14).

To find starting values for the optimization performed by mle2 we calculated the log-likelihood function for one million combinations of parameters values in a regular reticulate within reasonable ranges. The 100 sets of parameters that were local minima and with highest log-likelihood were used as starting values for the computational minimization.

The confidence intervals for the expected number of cases and frequency were estimated from 20000 parametric bootstrap samples assuming that the estimated parameters follow a multivariate normal distribution. The parameters of these multivariate distributions were the estimated values and estimated variance-covariance matrix of the parameters. For each sampled combination of para 2.5% and 95% quantiles of the distribution of bootstrapped expected values.

**D. Sensitivity analysis.** The model fitting assumed a constant infection hospitalization rate (IHR, parameter $\sigma$) for each age group over time for both variants. Recent evidence suggests that prior SARS-CoV-2 infection protects most individuals against reinfection (15), so reinfections might have lower IHR. Because the pathogenicity of the variant P.1 is unknown, the model fitting was repeated assuming that the odds ratio of the IHR in each age class for P.1 infections compared to wild-type variant infections (SA1) is a free parameter. Moreover, as the collapse of Manaus health system hindered hospitalizations of new severe cases and may have affected case recording in surveillance databases, the model fitting was repeated considering only the period prior to the collapse (10-January-2021) (SA2).
